# The miniECG: Enabling interpretable detection of amplitude and intraventricular conduction ECG-abnormalities with a novel ECG device

**DOI:** 10.1101/2025.06.03.25328866

**Authors:** Johanneke E. ten Broeke, Alejandra Zepeda-Echavarria, Rutger R. van de Leur, Melle Vessies, Manon G. van der Meer, René van Es, Rutger J. Hassink, Thierry X. Wildbergh, Rien van der Zee, Yaowen Zhang, Tjitske Heida, Joris E.N. Jaspers, Pieter A. Doevendans, Bram van Es

## Abstract

**Background:** The miniECG, a smartphone-sized, multi-lead device, offers a simple and fast alternative to the 12-lead ECG. We aimed to demonstrate the potential of the miniECG to record and detect amplitude and intraventricular conduction ECG abnormalities via interpretable models.

**Methods:** A miniECG was captured for patients undergoing a conventional 12-lead ECG in the University Medical Centre Utrecht. MiniECGs of patients with normal ECGs (controls), amplitude abnormalities (low QRS voltage (Microvoltage) and left ventricular hypertrophy (LVH)), conduction abnormalities (left or right bundle branch blocks (LBBB, RBBB), left anterior fascicular block (LAFB), bifascicular block (BfB)) were selected. Standard ECG-features were used as input for decision trees (DTs) for binary (normal vs abnormal) or multiclass classification, employing 10-fold stratified cross-validation.

**Results:** 1717 patients were included. For binary classification of conduction abnormalities, the decision tree showed an AUROC of 0.97±0.01 (NPV 0.95±0.03, sensitivity 0.95±0.02). For multiclass classification, AUROCs were 0.95±0.02 (BfB), 0.85±0.03 (LAFB), 0.87±0.05 (LBBB), 0.94±0.04 (RBBB) and 0.93±0.03 (controls). For binary classification of amplitude abnormalities, the AUROC was 0.76±0.04 (NPV 0.83± 0.05, sensitivity 0.84±0.06). For multiclass classification, the AUROCs were 0.80 ± 0.06 (LVH), 0.78±0.05 (microvoltage) and 0.76±0.05 (controls).

**Conclusion:** This is the first study to detect ECG abnormalities in amplitude and intraventricular conduction, with a four precordial electrodes set-up. DTs based on miniECG features offer an interpretable detection method with a performance that was comparable to other less interpretable models. Before clinical implementation, further research is necessary to optimize DT-structures and analyze abnormalities beyond the current study.

**Author Summary:** Electrocardiograms (ECGs) are vital for detecting abnormal electrical heart signals, but traditional 12-lead ECGs are often limited to in-hospital settings due to complexity. Our study introduces the miniECG, a smartphone-size device with four electrodes to be placed on the chest, that records eight leads. We evaluated its ability to detect amplitude and conduction cardiac abnormalities using interpretable machine learning models. In a hospital-based sample of over 1,700 patients, we trained decision trees (DTs) on standard ECG features to classify abnormalities. The models demonstrated accurate diagnostic potential, particularly for conduction abnormalities, while maintaining transparency and clinical interpretability. Further optimization is needed, but this tool could enable broader ECG acquisition and easier detection of cardiac abnormalities.

## Introduction

The electrocardiogram (ECG) represents the heart’s electrical activity and serves as the initial diagnostic tool for a wide range of electrophysiological abnormalities such as myocardial infarctions, arrhythmias, and conduction abnormalities. (1) These abnormalities can for example lead to cardiac ischemia, heart failure, or clinical symptoms such as palpitations, and should thus be detected to timely intervene (2–4). The 12-lead ECG is considered the gold standard to measure ECGs, as it provides a view of the heart’s electrical activity in twelve directions by placing electrodes on the limbs (4) and chest wall (6). (5) Annually, over 300 million 12-lead ECGs are measured worldwide, although the complexity involved in both the acquisition and interpretation have limited its utility mainly to hospital settings. (6) Due to the complexity of a 12-lead ECG measurement, only a selected pool of clinicians is trained to acquire them, which is inefficient and burdens clinical workflows. Easier methods to acquire ECGs have been developed, such as smartwatches or handheld devices. These devices are typically intended to detect rhythm disorders, such as atrial fibrillation (AF). (7,8) Less attention has been given to the detection of ECG-abnormalities beyond rhythm disorders, which requires multi-lead analysis. Examples include intraventricular conduction disorders, such as left bundle branch block (LBBB), which may lead to dyssynchronous contraction of the heart, reduced pump function and eventually heart failure. (3) Another example is low voltage ECG (Microvoltage), which arises when the generation or transmission of electrical signals to the electrodes is compromised, for example because of amyloidosis, pericardial effusion or obesity. (9) In contrast, left ventricular hypertrophy (LVH) is associated with high voltage ECGs, caused by increased thickness of the left ventricular wall. (10) Despite the clinical significance of detecting these ECG-abnormalities, there is a lack of faster and simpler alternatives to the traditional 12-lead ECG that can reliably identify such conditions.

The University Medical Centre Utrecht (UMCU) has introduced a mobile, smartphone-sized, handheld device capable of recording 8-lead ECGs using four precordial electrodes: the miniECG, based on a patented proof of principle of a handheld device with four adjustable electrodes.(11) The miniECG is placed on the chest (Fig 1) and captures a 10-second ECG in a simple and easy-to-use manner. For detection of ECG-abnormalities using the 12-lead ECG, extensive interpretation criteria regarding voltage, morphology or conduction times exist, applied by trained healthcare professionals to diagnose ECG-abnormalities. Because the miniECG’s electrodes are positioned differently than those of the 12-lead ECG, no established interpretation criteria exist for the miniECG. Therefore, it is necessary to investigate whether miniECG recordings can reveal electrophysiological abnormalities.

**Fig 1.**
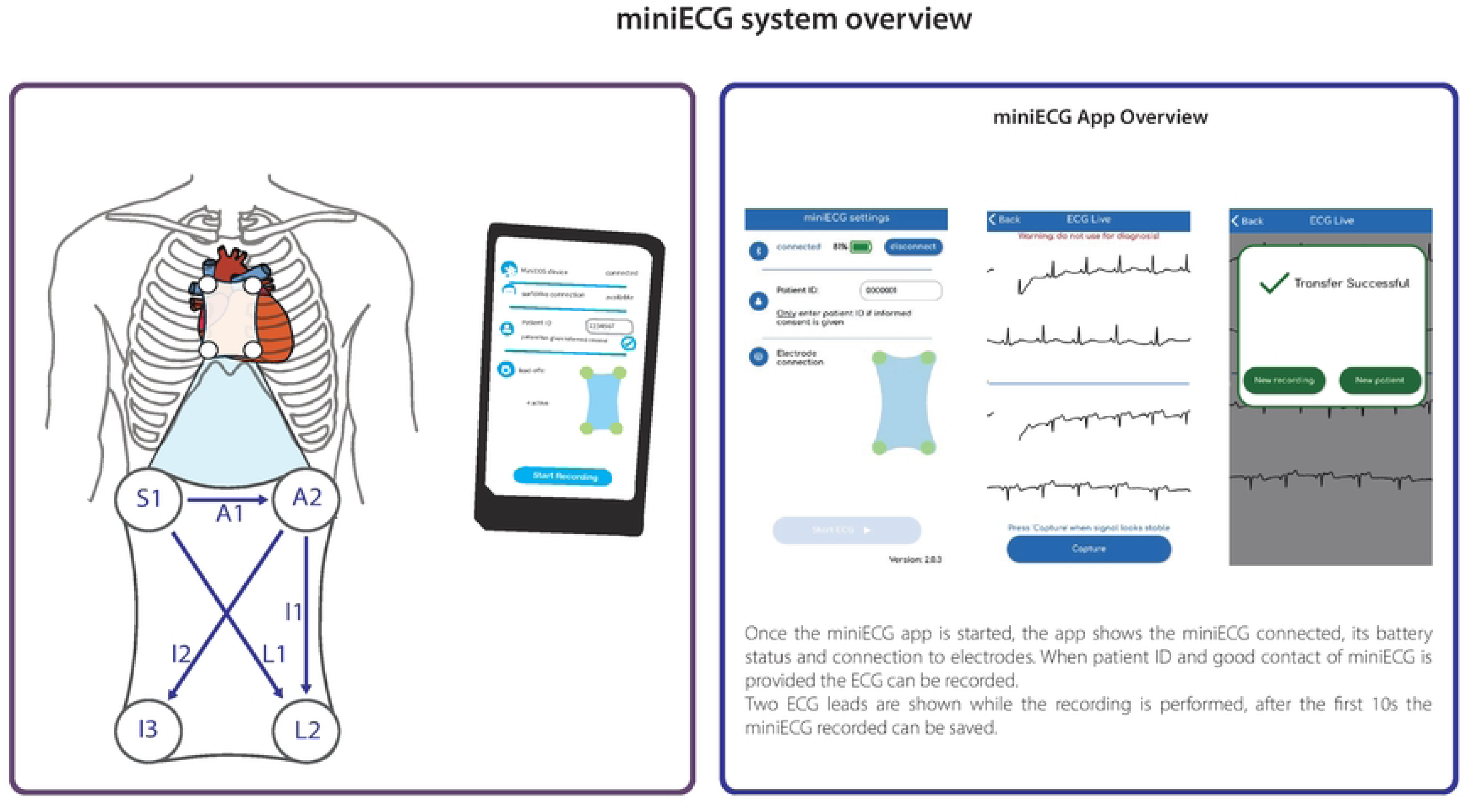
miniECG system overview, showing location of device over chest of patient at 2nd Intercoastal space, the recorded leads and overview of the app screens during use. The miniECG has four stainless steel electrodes that record eight ECG leads, four of them are bipolar leads (A1, L1, I1, and I2) and four unipolar channels (S1, A2, L2, and I3).

In previous studies, we showed the miniECG’s ability to capture ST-segment changes in the presence of coronary occlusions, demonstrating its potential for infarct detection. (12,13) In the present study, we investigate the miniECG’s ability to detect ECG-abnormalities - intraventricular conduction abnormalities and amplitude abnormalities-in hospital settings. This is a first attempt to assess such heart conditions while using the miniECG entailing a different placement than the 12-lead ECG. We prioritize interpretability of the detection methods, to provide clinicians with insights into how these abnormalities manifest on the miniECG. As we envision the miniECG serving as a screening tool prior to a 12-lead ECG, our focus was on minimizing false negatives to reduce the risk of missed diagnosis.

## Methods

We conducted a single-center study at the cardiology department and outpatient clinic of the University Medical Center Utrecht, Utrecht, the Netherlands (UMCU), between April 2022 and December 2023. Patients who underwent a 12-lead ECG (age >18 years, able to provide informed consent) were included. A 10 sec. miniECG was measured within a maximum of 10 minutes before or after the 12-lead ECG recording. Patients with anatomical restrictions (recent sternotomy or physical restrictions not allowing full contact with the miniECG) were excluded. The miniECG was placed over the sternal midline of patients; the two upper electrodes were placed at the second intercostal spaces. The miniECG with its four electrodes records eight ECG leads, four of them are bipolar leads (A1, L1, I1, and I2) and four unipolar channels (S1, A2, L2, and I3).

### Patient selection

Selection of patients was based on the automatic diagnosis of the 12-lead ECG derived by GE-Marquette 12SL ECG analysis program (GE Healthcare, Milwaukee, WI) and the physicians’ confirmation of diagnosis (if available). (14) The GE’s diagnosis criteria can be found in supplementary material 1. We selected patients with low voltage ECGs (referred to as Microvoltage), LVH and intraventricular conduction disorders (IVCD) including left bundle branch block (LBBB), right bundle branch block (RBBB), left anterior fascicular block (LAFB), and bifascicular block (BfB). We created two subgroups: the ‘conduction abnormalities’ group, including patients with LBBB, RBBB, LAFB and BfB, and the ‘amplitude abnormalities’ group, consisting of patients with LVH and Microvoltage. Each subgroup was compared to the control group, including patients selected solely on the absence of ECG abnormalities.

### MiniECG feature calculation

Once ECGs were recorded with the miniECG, postprocessing was performed. The device recorded 10 sec ECGs sampled at 250 Hz with a band-pass filter at 0.67–100 Hz and a 50 Hz notch filter.

For our analysis, standard ECG-features were calculated on the miniECGs as input for the DTs. MiniECGs were segmented to identify onsets and offsets of ECG-waves and to classify the QRS-complexes. A list of all possible labels is presented in supplementary material 2. Wave and noise segmentation was achieved using a previously published model that was a combination of 1) a widely adopted deep learning model, the deeplabV3, with 2) a convolutional neural network inspired by Hannun et al.. (15,16) This segmentation model was trained on 1937 physician annotated 12-lead ECGs obtained during routine care at the UMC Utrecht. [15-17].

Onsets and offsets of miniECG waves were determined identical across the eight leads, while noise detection was conducted separately per lead. Based on the classification of the QRS-complexes, the most common beats were selected. Beats containing noise or incomplete beats at the beginning or end of the recording were removed. For each remaining beat, the ECG-features depicted in figure 2 were determined. For the continuous features, the mean value over the beats was calculated.

**Fig 2.**
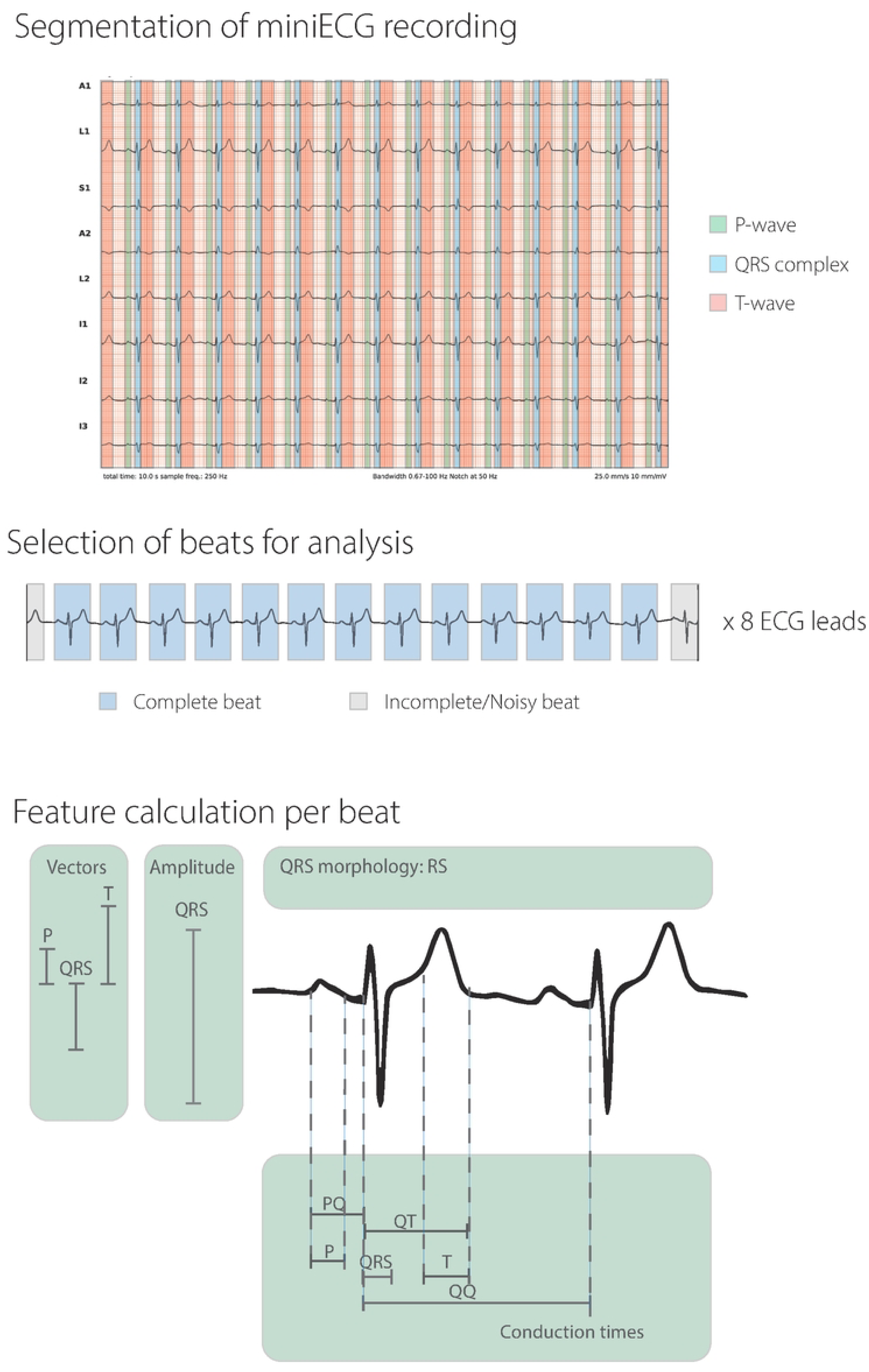
ECG analysis process, from segmentation of ECG waves, selection of beats for analysis and feature calculation per beat.

### Classification of miniECGs

Decision trees (DT) were constructed, trained and tested to perform the following classification tasks:

1. Binary classification for differentiating between patients with any conduction abnormality and controls (any conduction abnormality vs controls)
2. Binary classification for differentiating between patients with any amplitude abnormality and controls (any amplitude abnormality vs controls)
3. Multiclass classification for differentiating between the different conduction abnormalities and the control group (LAFB vs BfB vs RBBB vs LBBB vs controls)
4. Multiclass classification for differentiating between the amplitude abnormalities and the control group (Microvoltage vs LVH vs controls)

For amplitude abnormalities, the input features included the amplitude, vector, and morphologies of QRS complexes for each lead. For conduction abnormalities, the same features were used together with P- and T-vectors for each lead and the QRS-duration. For each classification task, a DT was constructed with gini impurity as splitting criterion. To balance usability and model performance, we limited the tree depth to six layers. We used balanced class weights, to automatically adjust the weights of each class, assigning higher weights to smaller classes to prevent model bias toward the more frequent classes.

For amplitude and conduction abnormalities, a binary and multiclass standard DT were constructed and for conduction abnormalities, an additional customized DT was constructed as well. This customized DT also had a maximum depth of 6 layers, but with a manually defined root node and one of its child nodes: the first split occurred at QRS-duration > 110 ms, followed by a second split at QRS-duration > 120 ms. These splits are based on guidelines for diagnosing IVCDs: in patients with a BBB, the QRS-duration exceeds 120 ms, for LAFB patients it’s below 120 ms, and a physiological duration is below 110 ms. (17,18)

The DTs’ performances were compared to that of XGBoost (XGB) and logistic regression (LR) to assess whether they achieve comparable performance to other machine learning methods that are commonly used for classification problems based on extracted features 10 fold stratified cross-validation was applied with 10 repetitions to both training and test set. We calibrated the model probabilities with an unweighted mean of sigmoid regressors based on the test-splits.

We compared the ROC-curves, calibration curves and net benefit curves of the DTs to those of LR and XGB. Net benefit curves were created to evaluate whether the models provide a higher clinical value compared to either classifying all ECGs as abnormal or classifying none as abnormal. (19–21). Mean and standard deviation were calculated over all folds and repetitions.

For the DTs, standard metrics (accuracy, F1-score, specificity, sensitivity, negative predictive value, positive predictive value) were calculated for a chosen probability threshold. Probability thresholds were manually chosen in such a way that NPV and sensitivity exceeded specificity where possible, to minimize the number of false negatives and avoid missing patients with abnormalities.

## Results

### Patient characteristics

Between 20 April 2022 and 5 December 2023, a miniECG was recorded for 7816 patients visiting the outpatient clinic. We selected 1717 unique patient recordings with intraventricular conduction and amplitude abnormalities on 12-lead ECG (Table 1). Of the diagnoses made by Marquette ™ (GE-algorithm), 19% were confirmed by a physician. There were 47 patients with overlapping abnormalities (21 LAFB + LVH, 11 BfB + LVH, 6 Microvoltage + LAFB, 3 Microvoltage + BfB, 3 Microvoltage + RBBB, 2 RBBB + LVH and 1 LBBB + LVH).

**Table 1.** Overview of the amount of included patients with their age and sex. *SD = standard deviation. Microvoltage = low (μV) voltage ECG. LVH = left ventricular hypertrophia. LBBB = left bundle branch block. RBBB = right bundle branch block. BfB = bifascicular block. LAFB = left anterior fascicular block*.

| Diagnosis | Amount<br>n (% female) | Age<br>mean (SD) |
| --- | --- | --- |
| All unique patients | 1717 (40%) | 61 ($\pm 16$) |
| Controls | 649 (49%) | 58 ($\pm 14$) |
| Microvoltage | 215 (57%) | 61 ($\pm 14$) |
| LVH | 198 (36%) | 64 ($\pm 17$) |
| LBBB | 150 (32%) | 68 ($\pm 13$) |
| RBBB | 207 (32%) | 61 ($\pm 18$) |
| BfB | 185 (19%) | 61 ($\pm 18$) |
| LAFB | 160 (29%) | 63 ($\pm 16$) |

### Architecture of the decision trees

The decision trees (DTs), constructed for each classification task, are accessible through GitHub at https://github.com/UPOD-datascience/TreeBuilder, where the split-criteria are shown together with their influence on the model probability and the actual coverage of each diagnosis in the dataset. To illustrate how the DTs can be applied, we provided an example path for LBBB (Fig 3) and for the other abnormalities in supplementary material 3. It is important to note that these paths are not the only route for that abnormality, and some miniECGs of patients with other abnormalities may meet the same criteria. In supplementary material 3, we explained the multiclass DTs in more detail.

**Fig 3.**
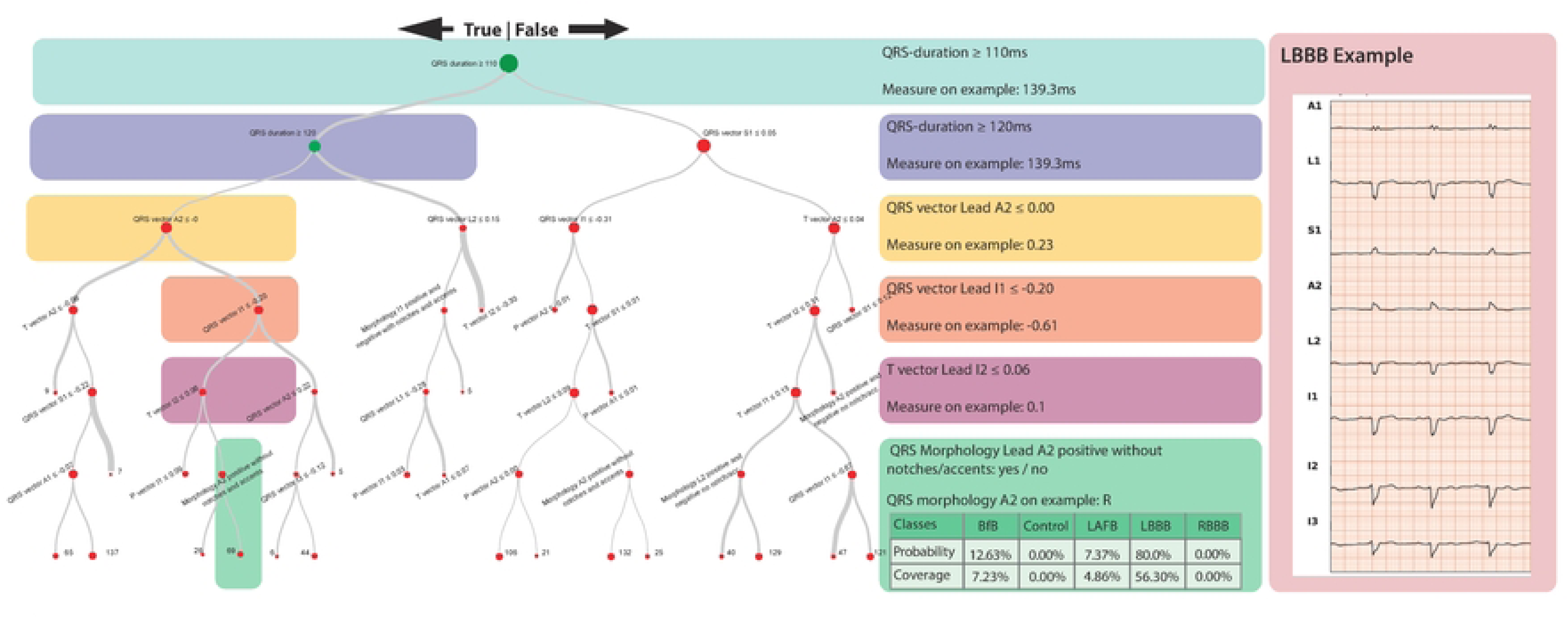
On the right (in orange), the first three beats of a miniECG of a patient with an LBBB are shown. The patient’s path through the DT is highlighted in colors. At each node, the splitting criterion consists of a specific feature and a cut-off value. If the patient meets this criterion, the path continues to the left child node; otherwise, it proceeds to the right. The probability for LBBB after the final split is 80%. In our dataset, 56% of all patients with an LBBB meet the same criteria and arrive in this node. It is highly probable that this patient has an LBBB, with a smaller chance of BfB or lafb. There is a 0% probability and coverage for this patient being a control, indicating the need for a referral to a 12-lead ECG to confirm the LBBB diagnosis.

### Performance of the decision trees compared to baseline models

#### Binary classification

In Fig 4 and Table 2, the ROC-curves for all classification tasks are shown. For binary classification of conduction abnormalities, the AUROCs were 0.94 ± 0.02 and 0.97±0.01 for the DT and the customized DT, respectively. The customized DT reaches the same AUROC as the extreme gradient boosting (XGBoost) model, whereas the linear regression (LR) shows a slight decrease in AUROC (0.93±0.02). For binary classification of amplitude abnormalities, the AUROC for the DT was 0.76 ±0.04, for the XGB 0.82±0.04 and for the LR 0.81±0.05.

**Fig 4.**
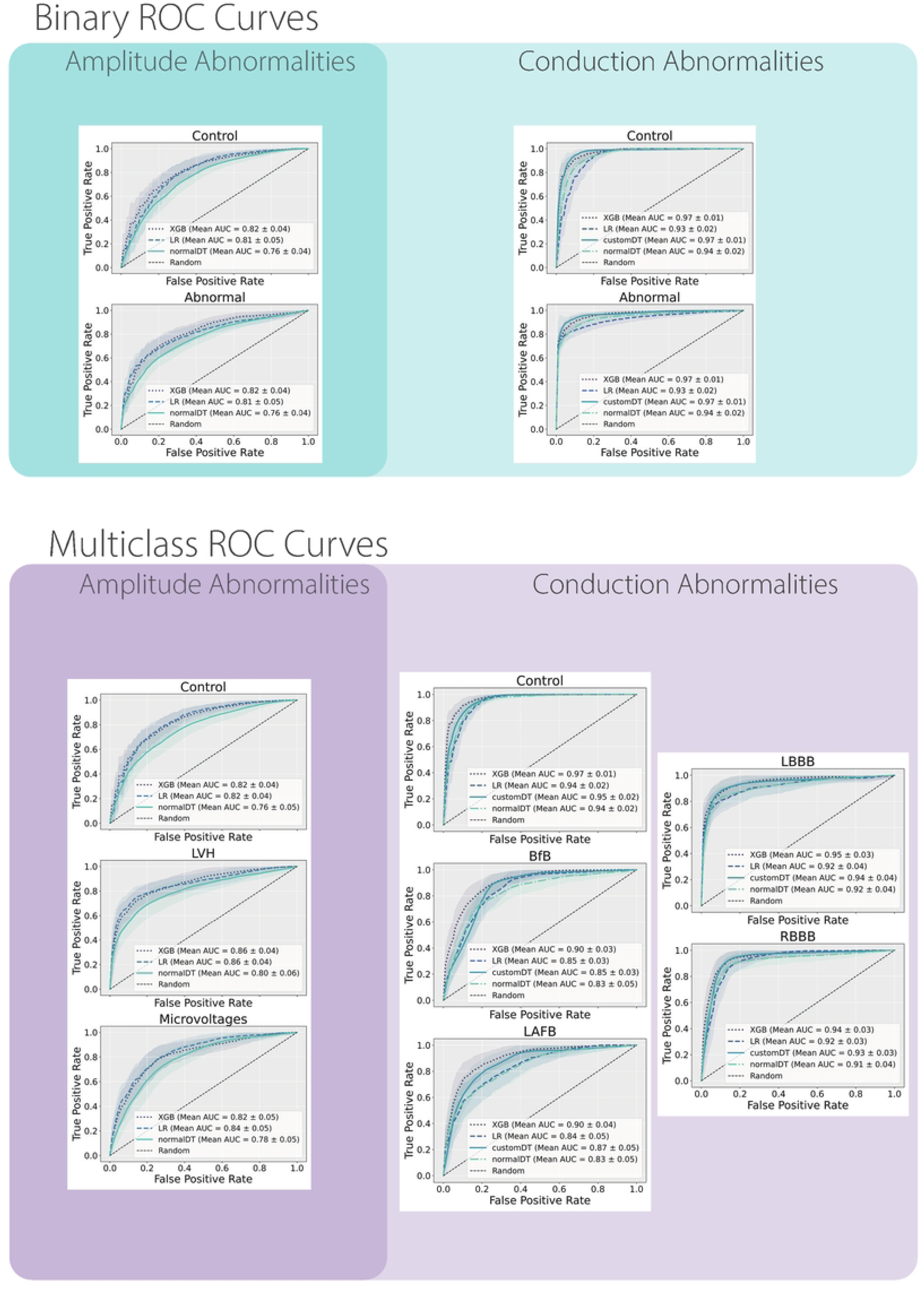
ROC-curves for conduction abnormalities (left) or amplitude abnormalities (right), for binary classification (top four figures) and multiclass classification. ROCs are shown for all models.

**Table 2.** AUCs for binary and multiclass classification for amplitude and conduction abnormalities. Normal DT: Normal Decision Tree, XGB: XGBoost, LR: Linear Regression, Custom DT: Custom Decision Tree

| Binary Classification AUC-ROC |  |  |  |
| --- | --- | --- | --- |
| Category | Diagnosis | Model | Mean AUC (SD) |
| Amplitude | Control | Normal DT | 0.76 (0.04) |
|  |  | XGB | 0.82 (0.04) |
|  |  | LR | 0.81 (0.05) |
|  | Abnormal | Normal DT | 0.76 (0.04) |
|  |  | XGB | 0.82 (0.04) |
|  |  | LR | 0.81 (0.05) |
| Conduction | Control | Custom DT | 0.97 (0.01) |
|  |  | Normal DT | 0.94 (0.02) |
|  |  | XGB | 0.97 (0.01) |
|  |  | LR | 0.93 (0.02) |
|  | Abnormal | Custom DT | 0.97 (0.01) |
|  |  | Normal DT | 0.94 (0.02) |
|  |  | XGB | 0.97 (0.01) |
|  |  | LR | 0.93 (0.02) |
| Multiclass Classification AUC-ROC |  |  |  |
| Category | Diagnosis | Model | Mean AUC (SD) |
| Amplitude | Control | Normal DT | 0.76 (0.05) |
|  |  | XGB | 0.82 (0.04) |
|  |  | LR | 0.82 (0.04) |
|  | LVH | Normal DT | 0.80 (0.06) |
|  |  | XGB | 0.86 (0.04) |
|  |  | LR | 0.86 (0.04) |
|  | Microvoltage | Normal DT | 0.78 (0.05) |
|  |  | XGB | 0.82 (0.05) |
|  |  | LR | 0.84 (0.05) |
| Conduction | Control | Custom DT | 0.95 (0.02) |
|  |  | Normal DT | 0.94 (0.02) |
|  |  | XGB | 0.97 (0.01) |
|  |  | LR | 0.94 (0.02) |
|  | BfB | Custom DT | 0.85 (0.03) |
|  |  | Normal DT | 0.83 (0.05) |
|  |  | XGB | 0.90 (0.03) |
|  |  | LR | 0.85 (0.03) |
|  | LAFB | Custom DT | 0.87 (0.05) |
|  |  | Normal DT | 0.83 (0.05) |
|  |  | XGB | 0.90 (0.04) |
|  |  | LR | 0.84 (0.05) |
|  | LBBB | Custom DT | 0.94 (0.04) |
|  |  | Normal DT | 0.92 (0.04) |
|  |  | XGB | 0.95 (0.03) |
|  |  | LR | 0.92 (0.04) |
|  | RBBB | Custom DT | 0.93 (0.03) |
|  |  | Normal DT | 0.91 (0.04) |
|  |  | XGB | 0.94 (0.03) |
|  |  | LR | 0.92 (0.03) |

The net benefit curves for binary classification in fig 5 show a net benefit for all binary models compared to either classifying all ECGs as abnormal or classifying none as abnormal. For the conduction abnormalities, the customized DT shows the highest net benefit, followed by the XGboost and the LR. For the amplitude abnormalities, the XGBoost shows the highest net benefit, followed by the LR and the DT. The XGBoost model has very poor calibration in all cases, as shown in the supplementary material 4, although this was monotonic and could therefore be mitigated with a monotonic regressor. The best out-of-the-box calibration for the conduction classification was obtained with the custom DT.

**Fig 5.**
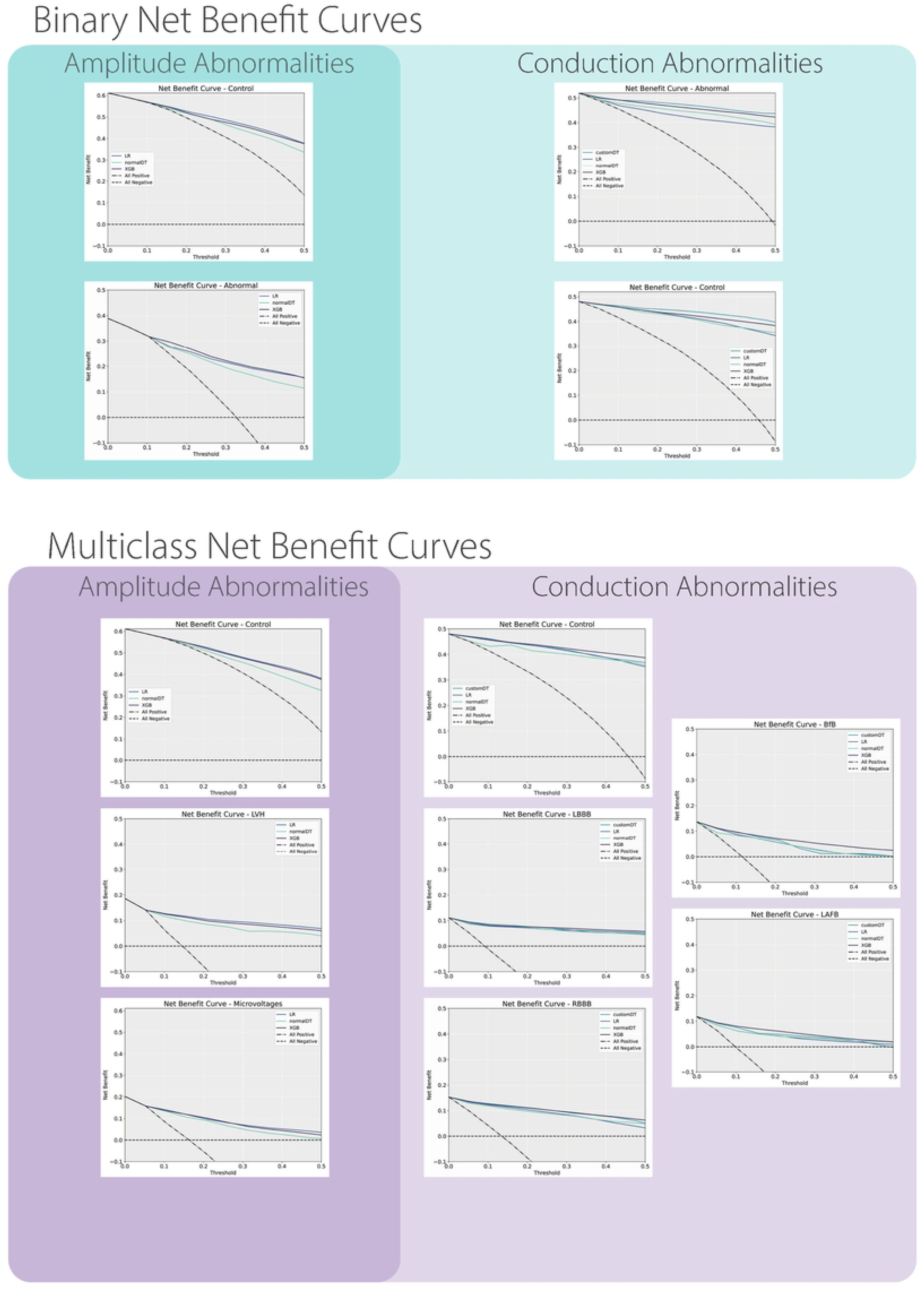
For binary classification of conduction abnormalities with the customized dt (left) or amplitude abnormalities (right) with the DT, the net benefit is visualized against the probability thresholds. Net benefit is calculated as: sensitivity × prevalence – (1 – specificity) × (1 – prevalence) × w where w is the odds at the threshold probability. Compared to classifying all patients as negative or positive (the black dotted lines), the introduction of all models show a net benefit, implying that intervening (e.g. by referring to a 12-lead ECG) based on the models leads to a higher net benefit than the alternative of intervening for all or none of the patients. For the conduction abnormalities, the customized DT shows the highest net benefit, followed by the XGboost and the LR. For the amplitude abnormalities, the XGBoost shows the highest net benefit, followed by the LR and the DT.

#### Multiclass classification

For multiclass classification of the conduction abnormalities, the customized DT shows AUROCs of 0.95±0.02, 0.85±0.03, 0.87±0.05, 0.94±0.04 and 0.93±0.03 for BfB, LAFB, LBBB, RBBB and the control group respectively. This is slightly higher compared to the AUROCs for the not-customized DT (0.95±0.02, 0.83±0.05, 0.83±0.05, 0.92±0.04 and 0.91±0.04 for BfB, LAFB, LBBB, RBBB and the control group respectively). Only XGBoost outperforms both DTs for all diagnoses (see Fig 4).

For classification of the amplitude abnormalities, the AUROCs of the DT were 0.80 ± 0.06, 0.78±0.05 and 0.76±0.05 for LVH, Microvoltage and the control group respectively. XGBoost and LR show similar AUROCs for the control group and LVH (0.82±0.04 for the controls and 0.86±0.04 for LVH). For the Microvoltage group, the LR outperforms the other models (AUROC of 0.84 ±0.05). The calibration curves can be found in supplementary material 4.

### Metrics for the decision trees

For the DTs (normal DT for amplitude abnormalities, customized DT for conduction abnormalities), the NPV, sensitivity and specificity for all threshold probabilities are shown in fig 6. For each classification task (one task per subfigure), one chosen threshold is displayed that ensures high NPV and sensitivity to minimize the number of false negatives. For these chosen thresholds, all metrics are shown in table 3 and 4.

**Fig 6.**
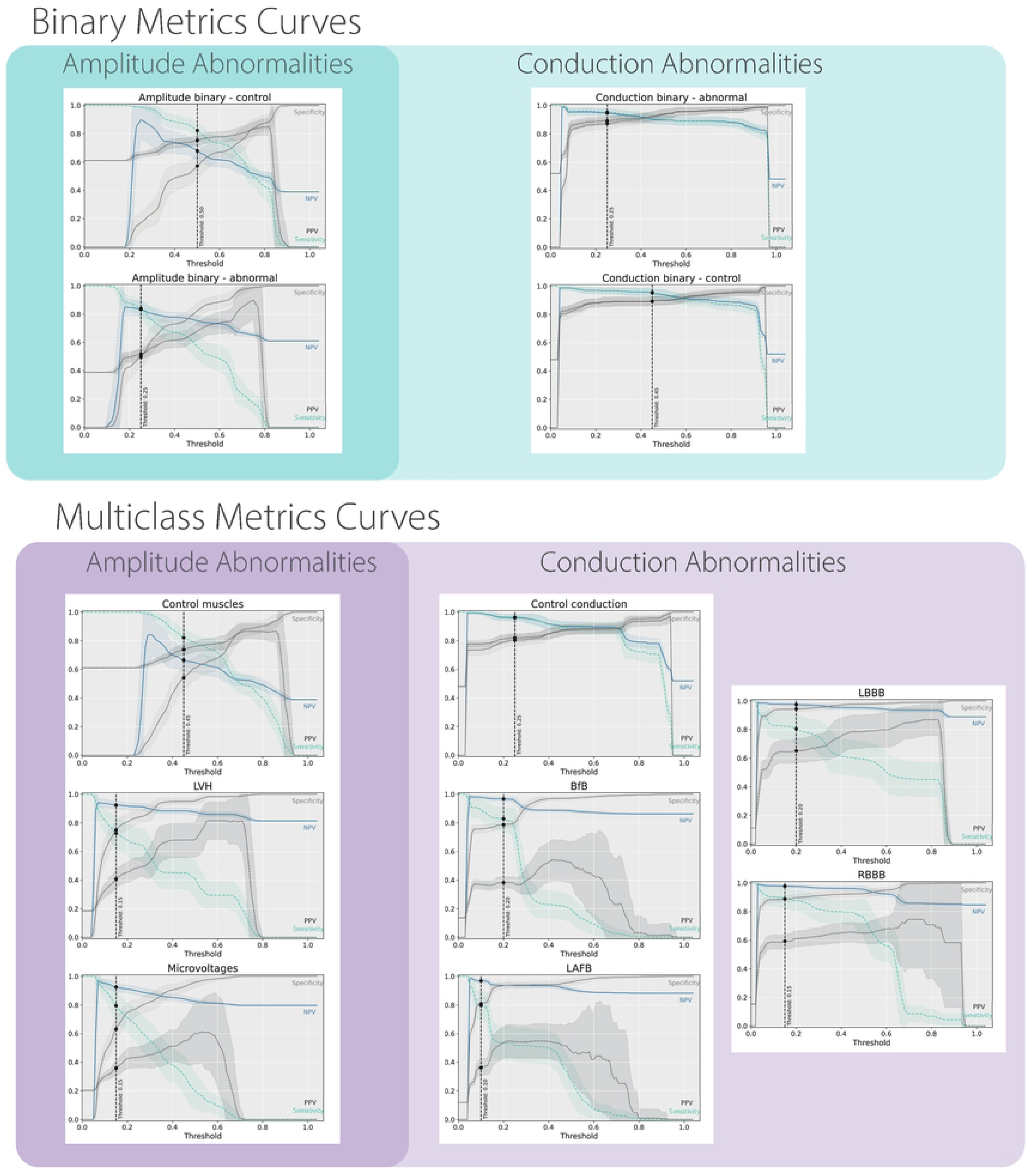
Metrics (specificity in grey, NPV in dark blue, sensitivity in light blue) against the thresholds for amplitude abnormalities (left), or conduction abnormalities (right), for binary classification (green) or multiclass classification (purple). The vertical black lines visualize the metrics for the chosen thresholds, for which the values can be seen in Tables 2 and 3.

**Table 3.** Metrics of decision trees for binary classification for amplitude and conduction abnormalities

| Category | Diagnosis | Threshold | NPV | Sensitivity | Specificity | PPV | Accuracy | F1-Score |
| --- | --- | --- | --- | --- | --- | --- | --- | --- |
| Amplitude | Control | 0.5 | 0.68<br>(0.06) | 0.82<br>(0.06) | 0.57<br>(0.09) | 0.75<br>(0.03) | 0.73<br>(0.04) | 0.79<br>(0.03) |
|  | Abnormal | 0.25 | 0.83<br>(0.05) | 0.84<br>(0.06) | 0.50<br>(0.07) | 0.52<br>(0.03) | 0.63<br>(0.04) | 0.64<br>(0.03) |
| Conduction | Control | 0.45 | 0.95<br>(0.03) | 0.95<br>(0.03) | 0.89<br>(0.03) | 0.89<br>(0.03) | 0.92<br>(0.02) | 0.92<br>(0.02) |
|  | Abnormal | 0.25 | 0.95<br>(0.03) | 0.95<br>(0.02) | 0.87<br>(0.04) | 0.89<br>(0.03) | 0.92<br>(0.02) | 0.92<br>(0.02) |

**Table 4.**
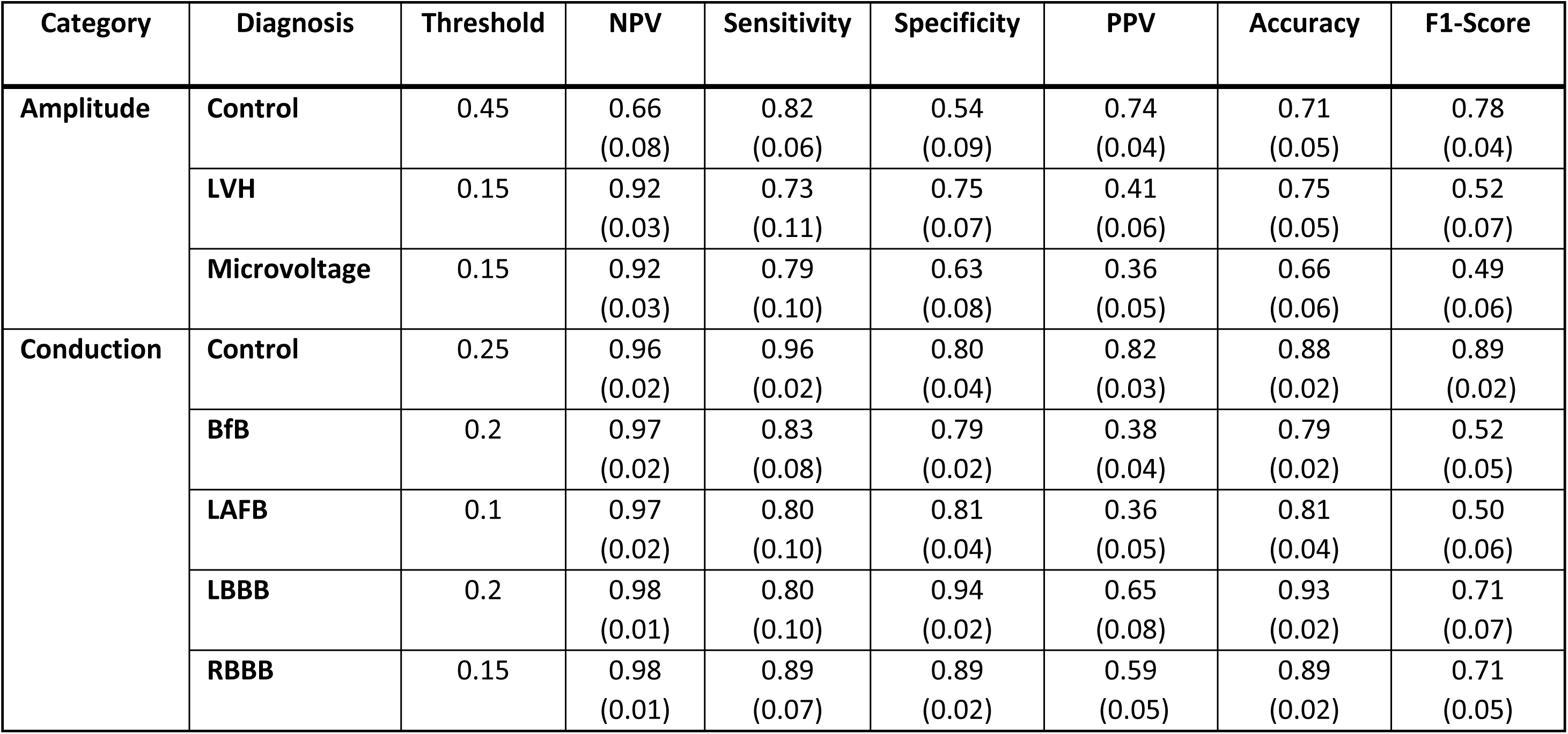
Metrics of decision trees for multiclass classification for amplitude and conduction abnormalities. *Microvoltage = low (μV) voltage ECG. LVH = left ventricular hypertrophia. LBBB = left bundle branch block. RBBB = right bundle branch block. BfB = bifascicular block. LAFB = left anterior fascicular block*.

## Discussion

We presented the first study to detect intraventricular conduction and amplitude ECG-abnormalities with a four precordial electrodes set-up. We believe that the miniECG’s multi-lead approach enables the diagnosis of a wider range of ECG-abnormalities compared to other 12-lead ECG alternatives. The DTs constructed in this study offer an interpretable method to detect conduction and amplitude abnormalities. They show comparable performance to less interpretable models (XGB and LR), are explainable and offer a net benefit. DTs’ AUROCs were higher for conduction than for amplitude abnormalities. For most abnormalities, NPV and sensitivity were already high, however they need to be improved to further eliminate false negatives before implementation into the hospital. In the following subsection, we will elaborate on the interpretation of the DTs’ split criteria, as they provide insight into how different ECG-abnormalities manifest on the miniECG and support a deeper understanding of the main results.

## Main results

### Intraventricular conduction abnormalities

For conduction abnormalities, the binary tree outperformed multiclass trees. This was expected, as it is simpler to differentiate abnormal from normal, than to identify specific abnormalities that manifest comparably on the ECG. Binary trees are thus preferred over multiclass trees for classifying conduction disorders in clinical practice, although multiclass trees provide a broader diagnostic scope, so their combined use is valuable. The customized DT, with preset QRS-duration criteria in the top nodes, outperformed the standard DT, making it the preferred option.

In the multiclass trees, QRS-duration is a key feature. The physiological QRS-duration is below 110 ms, patients with an LAFB have a QRS-duration below 120 ms while in patients with a BBB or BfB it exceeds 120 ms. For RBBB and LBBB, the DT’s performance is the highest. They can be differentiated from LAFB and controls based on QRS-duration, and from each other by using QRS-vectors in multiple leads, as the direction of the mean electrical activity, reflected in QRS-vector, depends on the location of the block. Differentiating RBBB from BfB is more challenging, as BfB is a combination of RBBB and either LAFB or left posterior fascicular block (LPFB), so the two manifest quite comparably on the ECG, which we also see on the miniECG: there is not a single tree path of miniECG-criteria that is met by RBBB-patients and not by BfB-patients, or vice-versa. This may explain the relatively low performance for BfB-detection. For LAFB, performance is also relatively low. While QRS-duration allows differentiation from other blocks, it does not distinguish it from controls. In LAFB patients, the abnormal path of the electrical conduction alters the heart axis, which can only be partially assessed with the miniECG as we measure only in the horizontal plane. This may explain the relatively low performance.

### Amplitude abnormalities

For amplitude abnormalities, the AUROCs for the multiclass trees were slightly higher than for the binary tree. This was anticipated, as in the Microvoltage group QRS-amplitude is lower than in controls whereas it is higher in LVH, making it easier to differentiate the Microvoltage and LVH groups from each other when evaluated as separate abnormalities, than to differentiate normal from abnormal. The multiclass trees would thus be preferred over the binary tree in clinical practice.

The amplitude DTs show lower performance compared to the conduction DTs. This may be explained by the miniECG’s electrode placement closer to each other compared to the 12-lead ECG reduces overall QRS voltages: differences between amplitude abnormalities and control may not be as distinguishable as expected.

### The miniECG in the context of established literature

Other alternatives for the 12-lead ECG, such as wrist watches, are primarily designed for detecting rhythm disorders, mostly AF. (8,22–26). In contrast, the multi-lead miniECG can identify other, more complex, cardiac abnormalities, which remains challenging for 1-lead devices.

The challenges in detecting heart abnormalities stem from the underlying detection principles. While rhythm abnormalities can be detected with a single-lead, this is not the case for other heart abnormalities. For example, Caillol *et al.* reported limitations when using a single-lead ECG for the detection of myocardial ischemia, as anterior infarctions may go undetected (27). Similarly, intraventricular conduction disorders require multi-lead analysis of waveform morphologies for diagnosis. Single-lead devices may not capture the full electrical activity of the heart, limiting their diagnostic utility in these cases.

The performance of single-lead and home-use devices in detecting other cardiac abnormalities beyond rhythm abnormalities has been explored by few studies. Nigolian et al. demonstrated high accuracy (0.96, 0.98, 0.98) for detecting LBBB, RBBB, and LVH using a single-lead handheld ECG device positioned in nine different locations, outperforming the miniECG (0.93, 0.89, 0.75)(28). However, their study had smaller sample sizes, resulting in wide 95% confidence intervals. In contrast, our study included additional abnormalities (BfB and LAFB), which may reduce accuracy but increase clinical relevance. Other studies using a single-lead device (Lead I) or a 12-lead belt reported perfect specificity and PPV but lower sensitivity (0.875 and 0.464, respectively)(29,30) Our study’s larger sample size (702 patients) yielded a sensitivity of 0.95, and the miniECG demonstrated balanced performance across sensitivity, specificity, PPV, and NPV, showing promise for accurate detection of atrioventricular conduction disorders. (27–29) The use of 1-lead ECG devices for detecting ECG-abnormalities remains challenging, as these conditions often require multi-lead recordings.(27)To compensate the missing leads, studies have shown alternative uses by proposing serial recordings performed over different points of the chest similarly to the 12-lead ECG (31–33). However the multi-lead serial recordings with single-lead devices can be cumbersome for patients, as they require precise placement, and are time consuming, while the recordings with the miniECG would allow to a multi-lead approach by placing the devices in a single place.

### Strengths and limitations

One of the strengths of our study is the large, hospital-based sample size (1717 patients), which supports robust performance evaluation of the miniECG beyond rhythm abnormalities. All ECG-diagnoses were represented by at least 150 patients each, enhancing both the reliability and broader applicability of the results. Another strength is the selection of abnormalities apart from rhythm disorders and beyond only complete BBBs, as before implementation in the hospital, it is necessary to reliably detect the full spectrum of ECG abnormalities with the miniECG. Besides, standard ECG-features were used as input for the DTs, allowing HCPs to interpret miniECG measurements based on features they are already accustomed to and facilitating easier integration into clinical practice.

The miniECG’s design with four precordial electrodes allows for quick and user-friendly acquisition of ECGs, with this study we showed its potential for screening purposes. Although the miniECG records 8 leads from just four chest electrodes, its novel electrode configuration allows for a compact and user-friendly design. However, this setup inherently limits recording to the horizontal plane, which may explain challenges in detecting certain abnormalities such as LAFB. Compared to the 12-lead ECG, where frontal and horizontal planes are both covered, the lack of limb lead equivalents limits the spatial perspective needed for some conduction abnormalities.

A limitation is the reliance on the GE-algorithm, lacking perfect labeling for some ECGs, as a large part was not confirmed by a physician. For the abnormalities for which the algorithm’s detection performance was reported (LBBB, RBBB, LVH), performance was good. (14,34) The algorithm has been validated in multiple clinical settings, adapted in clinical practice and is therefore the gold standard algorithm for automated ECG-diagnosis. Another limitation is the lack of focus on differences in age, sex or other clinical or demographical patient characteristics in this study. As the ECG is influenced by these factors, the lack of matching between patient groups based on these characteristics can decrease the model’s performance and generalizability. (34) Developing models tailored to specific age or sex groups may improve this. Moreover, body mass index (BMI) information was not available for our patients. In future research, the effect of BMI on the miniECG’s QRS-amplitude should be investigated to improve performance when detecting amplitude abnormalities. Furthermore, features were extracted using a segmentation model that was trained on 12-lead ECGs and not on miniECGs. This may introduce small errors in conduction times, although the segmentation is expected to be fairly accurate as fundamental characteristics of a miniECG and 12-lead ECG are comparable. Future validation of the segmentation model on miniECGs may improve the miniECG’s detection performance further. Finally, 10-fold cross validation was performed, resulting in multiple decision trees that were trained and tested, with the mean of the metrics calculated. The low standard deviation across all metrics implies consistent performance across the trees. However, the stability of the trees, regarding features and the cut-offs used for the splits requires further analysis. Future research should focus on combining these trees into a single, more robust decision tree.

### Future perspectives for clinical use

As a potential front-line screening tool within hospitals, the miniECG could help identify patients requiring follow-up with a 12-lead ECG, improving workflow efficiency and broadening access to ECG diagnostics. Studies suggest that the use of home monitoring devices can alleviate pressure on healthcare systems; however, current care models must be adapted to ensure that patient management remains effective. (35–37)

Before implementing the miniECG into clinical practice, the positioning of the device requires attention. It can be challenging to position the device consistently and correctly and mispositioning influences the ECG and features derived from it. In the presented study, miniECGs were acquired by trained HCPs so we expect the impact of positioning on the results to be minimal. However, the impact of the position on the chest should be investigated thoroughly, especially if patients use the device at home in the future. Besides, the lack of leads in the horizontal plane limits our ability to measure the heart’s electrical activity in all directions. Positioning the device in an extra position, for example lateral, may help acquire this extra information and improve detecting performance further.

## Conclusion

In conclusion, our study demonstrates the potential of the miniECG for the classification of intraventricular conduction and amplitude abnormalities. DTs offer an interpretable method for the detection of these abnormalities. They show a minimal decrease in performance compared to other – less interpretable – machine learning models. Future steps, such as optimizing the segmentation method, refining the DT-structure, and including patients with abnormalities beyond those of the current study, should be undertaken before implementation in clinical practice. After these steps, we believe the miniECG has the potential as a screening tool to evaluate who should undergo a 12-lead ECG, and expand the pool of clinicians able to acquire ECGs due to its user-friendly design consequently improving clinical workflow efficiency.

## Data Availability

The data that support the findings of this study are available on reasonable request to the corresponding author.

https://github.com/UPOD-datascience/TreeBuilder

## Acknowledgements

We kindly acknowledge all the clinical staff that supported the development and use of the miniECG at the UMC Utrecht Poli and Heart Function Departments.

## Supplements

**Supplement 1 :**
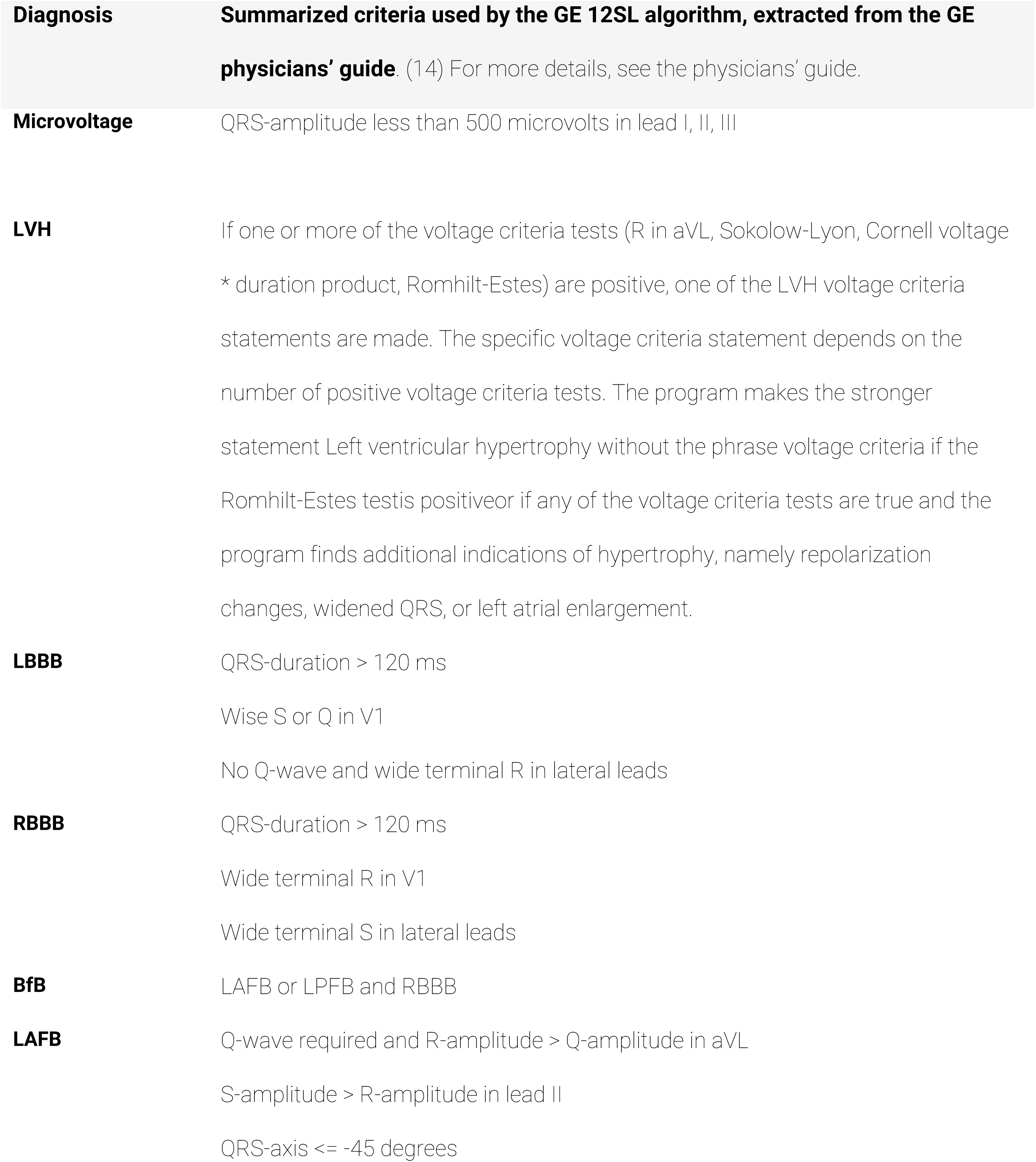
GE 12SL algorithm – diagnostic criteria used for patient selection.

| Diagnosis | Summarized criteria used by the GE 12SL algorithm, extracted from the GE physicians' guide. (14) For more details, see the physicians' guide. |
| --- | --- |
| **Microvoltage** | QRS-amplitude less than 500 microvolts in lead I, II, III |
| **LVH** | If one or more of the voltage criteria tests (R in aVL, Sokolow-Lyon, Cornell voltage * duration product, Romhilt-Estes) are positive, one of the LVH voltage criteria statements are made. The specific voltage criteria statement depends on the number of positive voltage criteria tests. The program makes the stronger statement Left ventricular hypertrophy without the phrase voltage criteria if the Romhilt-Estes testis positive or if any of the voltage criteria tests are true and the program finds additional indications of hypertrophy, namely repolarization changes, widened QRS, or left atrial enlargement. |
| **LBBB** | QRS-duration > 120 ms<br>Wise S or Q in V1<br>No Q-wave and wide terminal R in lateral leads |
| **RBBB** | QRS-duration > 120 ms<br>Wide terminal R in V1<br>Wide terminal S in lateral leads |
| **BfB** | LAFB or LPFB and RBBB |
| **LAFB** | Q-wave required and R-amplitude > Q-amplitude in aVL<br>S-amplitude > R-amplitude in lead II<br>QRS-axis <= -45 degrees |

**Supplement 2:** the labels used for segmenting miniECGs.

| P-wave | Qrs-complex | T-wave | Other |
| --- | --- | --- | --- |
| Sinus rhythm | Sinus rhythm | Normal T-wave | Noise |
| Premature atrial complex | Premature ventricular complex |  |  |
| Atrial pacing | Premature atrial complex |  |  |
| Atrial rhythm | Atrial fibrillation |  |  |
| No AV-conduction | Atrial flutter |  |  |
|  | Atrial rhythm |  |  |
|  | Nodal escape |  |  |
|  | Ventricular pacing |  |  |
|  | Atrial pacing |  |  |
|  | Fusion |  |  |
|  | Supraventricular tachycardia |  |  |
|  | Unknown |  |  |
|  | Ventricular tachycardia |  |  |
|  | Ventricular escape |  |  |

### Supplement 3: Explanation trees

#### LAFB/controls vs BfB/LBBB/RBBB

In the customized decision tree for conduction abnormalities, we started with the criterion QRS-duration < 110ms, which effectively differentiated between LAFB/controls and other abnormalities: 98% of the controls and 77% of LAFB patients met this criterion, while more than 90% of BfB, LBBB and RBBB patients did not meet the criterion.

#### LAFB vs controls

To further differentiate between LAFB patients and controls, the QRS-vector in lead S1 can be used: 53% of the control patients and only 6% of the LAFB patients have a vector below 0.05mV. Therefore, a QRS-duration below 110ms and a QRS-vector S1 < 0.05mV is most likely a control patient (probability of 94%). Controls and LAFB-patients with a QRS-vector exceeding 0.05mV (43% of control patients, 72% of LAFB-patients) show quite similar ECG-characteristics: in 40% of controls and 59% of LAFB-patients, the T-vector in lead A2 is below 0.04mV and the T-vector in lead I2 is below 0.31mV. The combination of an I1 T-vector exceeding 0.15 and a QRS-vector lead I1 below -0.67 is most efficient in differentiating further (4% of controls, 18% of LAFB-patients).

#### LBBB vs RBBB/BfB

Assessing the QRS-vector in lead A2 helps differentiating between LBBB and RBBB-patients: in 71% of the RBBB patients, this vector is below 0mV, whereas in 72% of the LBBB patients, it exceeds 0mV. It does not help to classify BfB patients: it is below 0mV for 44% of BfB-patients, and exceeds 0mV for 36%. So, an A2 QRS-vector vector exceeding 0mV is most likely a LBBB-patient, although in 36% of BfB patients and in 15% of RBBB patients, the QRS-vector in lead A2 also exceeds 0mV. To differentiate between RBBB/BfB-patients and LBBB-patients, the QRS-vector in lead I1 can be inspected: it is above –0.2mV in only 0.7% of the LBBB-patients and in 18% of the BfB-patients and 12% of RBBB-patients. Further differentiation between BfB-patients and RBBB-patients is not possible.

If the I1 QRS-vector is below 0.2mV (18% of BfB-patients, 3% RBBB, 71% LBBB patients), the T-vector in lead I2 helps differentiating further (exceeding 0.06mV in 7% of the BfB-patients and in 56% of LBBB patients). If the I2 T-vector is below 0.06mV, it’s not possible to further differentiate between LBBB and BfB.

#### BfB vs RBBB

For 44% of BfB patients and 71% of RBBB patients, the QRS-vector in lead A2 is below 0mV. There are no features in the DT that efficiently help to further differentiate between BfB and RBBB patients.

**See Figures in Supplementary Figures**

Supplement 4: Calibration curves

**See Figures in Supplementary Figures**

## Notes

### Competing Interest Statement

The authors have declared no competing interest.

### Funding Statement

Yes

### Author Declarations

The study is a non-WMO study approved by the institutional review board at the University Medical Center Utrecht.

